# COVID-19 in London, a Case Series Demonstrating Late Improvement in Survivors

**DOI:** 10.1101/2020.05.16.20103853

**Authors:** Ahmed Al-Hindawi, Jagdish Sokhi, Joshua Cuddihy, Christopher Lockie, Linsey Christie, Roger Davies, Suveer Singh, Marcela Vizcaychipi, Michelle Hayes, Alice Sisson, Richard Keays

## Abstract

**Objective:** To determine whether the trajectories of survivors and non-survivors are different in patients admitted to intensive care in London.

**Design:** In this case series of 15 survivors and 16 non-survivors, data from admission to discharge was collected and aligned to lowest PaO2/FiO2 ratio where aggregation and trends were demonstrated.

**Setting:** Single centre case-series in London, Intensive Care.

**Participants:** All non-survivors were included (n = 16). A biased set of survivors (n = 15) who were demonstrated an unexpected and rapid recovery after a prolonged period of mechanical ventilatory support.

**Results:** Respiratory failure trajectories of survivors and non-survivors were similar once aligned indicating, from a respiratory function perspective, it is difficult to identify survivors from non-survivors with some survivors improving late in their disease (day 20 – 30 from symptom onset)

Non-survivors are admitted earlier in their disease (p<0.05) and had worse organ failure requirements prior to the nadir of their respiratory funciton (p<0.05) compard to survivors.

**Conclusion:** Analysis of multiple factors fails to differentiate between survivors and non-survivors. Even when faced with multiorgan failure, perseverance until discharge must be advocated as late improvements do occur in survivors.

## Introduction

The SARS-COVID-2 virus giving rise to the syndrome of COVID-19 is a world-wide pandemic. Multiple case reports from China and America have highlighted the seriousness of the disease with increased mortality (49 – 88%), length of stay and time of presentation
^1^–^3^. In those publications, the median time from onset of symptoms to ICU admission was around 10 –11 days. The median time from illness onset to mechanical ventilation was 14.5 days, and the median time from illness onset to death was 18.5 days^1^. Zhou *et al* demonstrated that, in their retrospective cohort study, 32 Wuhan patients required invasive mechanical ventilation, of whom 31 (97%) died with Extracorporeal Membraneous Oxygenation (ECMO) was used in 3 patients who all died. The ICU length of stay in non-survivors was a median of 8 days (range 4–12 days). A more comprehensive analysis by the Chinese Centre for Disease Control and Prevention showed that the case fatality rate, across China, for patients admitted to intensive care was 49% (2087 admitted to ICU out of the 72,314 patients)^4^.

In New York, it was reported that the mortality in patients who received mechanical ventilation was 76.4% in 18–65-year olds and 97.2% in those over 65 years. This did not include the patients still remaining in hospital whose outcome was undetermined up to that point^2^. In Washington State, an ICU mortality of 67% has been reported whilst a further 24% of patients still remained on ICU with renal failure occurring more frequently in non-survivors^5^. None of the survivors had a serum ferritin greater than 2000 µg/L.

In the United Kingdom, as of the 8^th^ of May 2020, of the critical care outcomes reported (8250 patients), the mortality rate stands at just over 46.8%. The total number of patients admitted to ICU is 10758 and even if all the patients remaining in hospital were to survive the death rate would still be around 33%. Over 60% of these deaths occur within the first 10 days of admission to ICU and 92% of deaths have occurred by day 20. The median length of stay amongst non-survivors was 7 days (4 – 13 days). Amongst patients who only required advanced respiratory support, mortality was 43.9% but if renal failure supervened mortality increased to 65.4%^6^.

It has been advised that triaging is necessary in the context of overwhelmed medical services: J. Phua *et al* state to ‘Consider critical care triage to ration scarce resources in pandemic; prioritise patients who will benefit from ICU care’^7^. The concept of triaging during a pandemic and consideration of resource allocation has been implied at a UK National level, although there have been various iterations as time has gone by^8^.

The purpose of this paper is to describe the experience with COVID-19 patients mechanically ventilated on ICU who showed an unexpected and rapid recovery after a prolonged period of mechanical ventilatory support. Many of these patients required vasopressor infusions, multiple episodes of proning and some required renal replacement therapy culminating in a crude, unadjusted, mortality of 27%. We contrast the survivor trajectory with non-survivors demonstrating that the two groups are relatively indistinguishable until discharge.

## Methodology

The study was approved by the Chelsea and Westminster NHS Foundation Trust clinical governance team. As the report involves routinely collected non-identifiable clinical audit data, no ethical approval was required under the UK policy framework for health and social care.

Data collected SARS-CoV-2 positive patients addmitted to Chelsea and Westminster NHS Foudnation Trust Intensive Care Unit from 9^th^ of March till the 5^th^ of May 2020. All non-survivors were included in collection and analysis. A sample of survivors was chosen who had a prolonged course on ICU with features which pointed to a poor outcome but who went on to recover. This cohort of survivors thus represents patients who, given the current literature, withdrawal of care would have been considered.

Observational, pathological, and ventilatory values from patients were extracted from Cerner Electronic Patient Record (EPR) and analysed in R 3.6. The lowest PaO_2_/FiO_2_ (PF) ratio of an arterial sample on each day was recorded.

To facilitate aggregation and identification of trends, patients’ symptom days were aligned such that the lowest PF ratio on ICU constituted Day 0. Negative day numbers (as seen in Figure 1 and Figure 3) thus represent days patients were on ICU prior to the lowest PF ratio. In Figure 2, day 0 is defined as the first day of symptom onset; patient trajectories commence from ICU admission.

**Figure 1.**
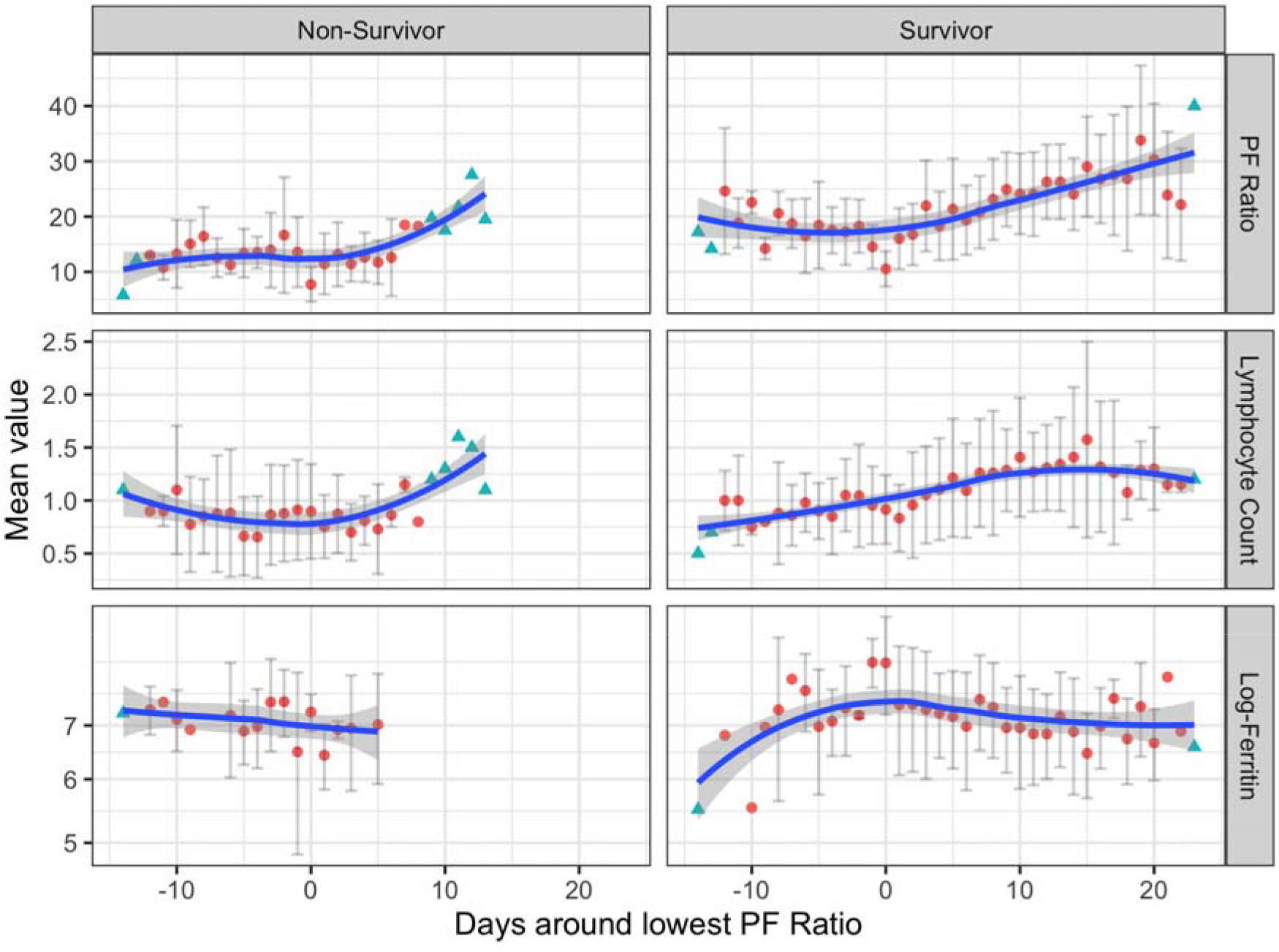
Three plots demonstrating recovery of lymphocytes proceeding the nadir of P/F ratio concurrent with the peak of ferritin level. Data aggregated by aligning patients to the nadir PF ratio. Blue triangles represent data points which were informed with only 1 patient and thus do not have error bars. Error bars represent 1 standard deviation of values for that day. All values are used for LOESS trend line regression.

**Figure 2.**
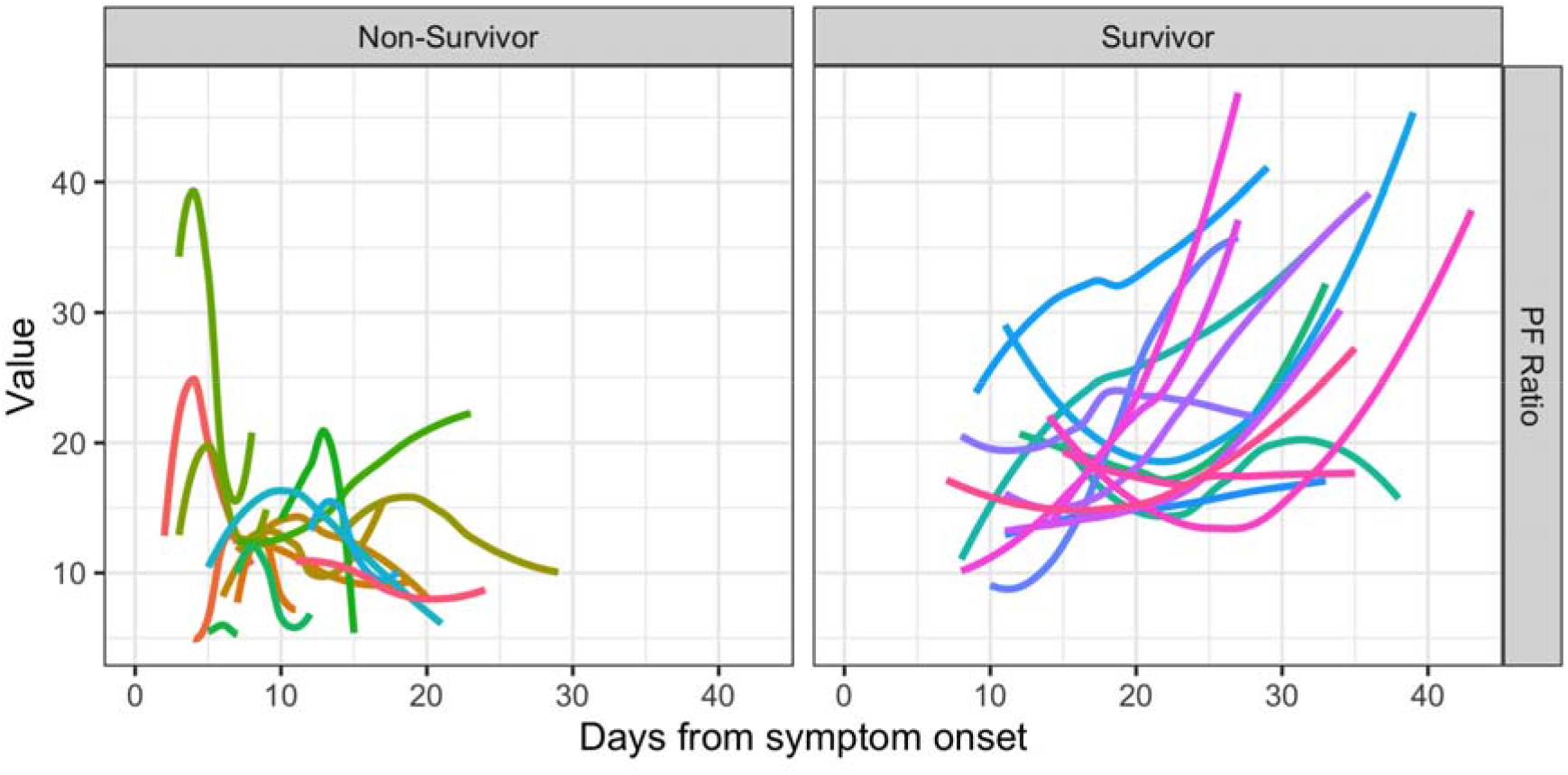
LOESS trend line for each patient per variable from date of onset of symptoms. Data collection commenced on admission to ICU. Day 0 here is the day the patient’s symptoms began and thus the period from symptoms commence till ICU admission lacks data.

**Figure 3.**
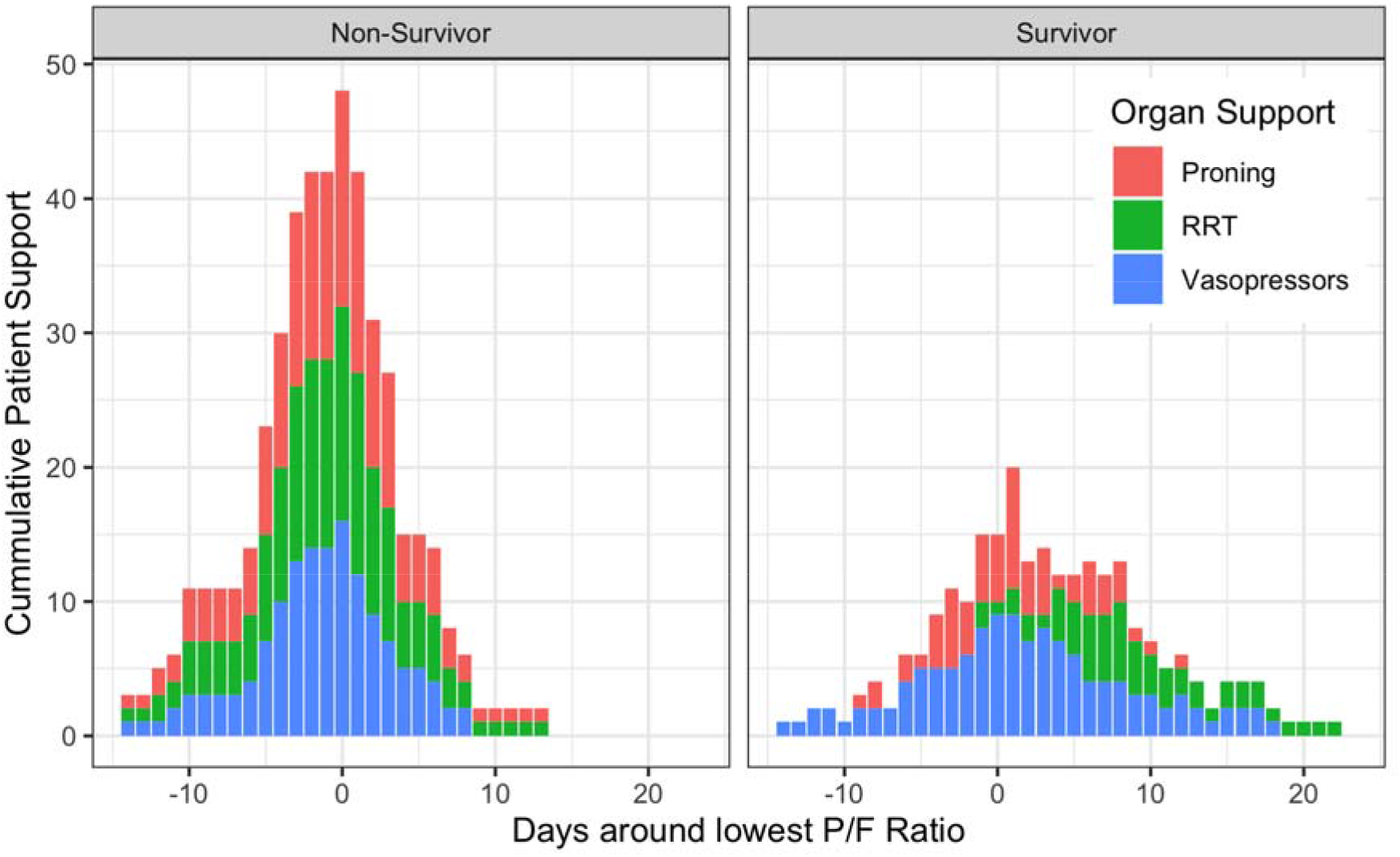
P/F Organ support required for patients in ICU over the course of their stay. The day coinciding with the lowest P/F ratio has significantly higher requirements for organ support for both groups of patients. Of note, vasopressor use reduces but renal replacement therapy and proning continue to stay elevated till circa day 10 and then decline rapidly. Some survivors require ongoing renal replacement therapy. Non-survivors have a statistically significant (P < 0.05) distribution of requiring renal replacement therapy earlier compared to survivors.

As this is a descriptive case-series, three patient-based variables were focused on due to their interest in COVID-19 literature; namely the PF ratio; lymphocyte count and ferritin. Due to the magnitude of ferritin values across patients, the natural-log is used.

For organ support variables, proning, vasopressors and renal replacement therapy were chosen. Vasopressor use was deemed if any of noradrenaline, vasopressin or metaraminol were delivered during that day. Proning episodes were extract from free text entries in Cerner EPR.

For visualisation, three figures were made. Figure 1 and Figure 2 have trend lines, grouped by mortality, that were calculated according to LOcally Estimated Scatterplot Smoothing (LOESS) with a parameterised span of 1; the grey area demonstrating 1 standard deviation.

Figure 3 demonstrates organ support required over the course of the patient’s stay in ICU gouped by mortality.

Distributions in Figure 3 were tested for normality using Shaprio-Wilk test followed by an unpaired T-Test.

## Results

Survivors (n=15) and non-survivors (n=16) were analysed. Figure 1 demonstrates that, when patients are aligned from their lowest PF ratio day, there are similar trajectories in the survivors and non-survivors. Non-survivors start with a lower PF ratio than survivors, but PF nadir alone does not determine death as ratios recover in both groups. Survivor PF ratios improve to a greater extent than that of non-survivors. Lymphocytes also recover with little to be gleaned from absolute values whereas there appears a trend in ferritin levels that differentiate survivors from non-suvivors; ferritin levels are initially higher in non-survivors and plateau unlike survivors whose values recover from their zenith.

Figure 2 demonstrates that non-survivors are admitted to ICU earlier than the survivors from the day their symptoms start (unpaired T-Test; t = –3.7137, p < 0.001). Similarly, to Figure 1, the trend of lower PF ratio in non-suvivors compared to survivors can be seen.

Figure 3 demonstrates organ support as defined by the use of vasopressors, renal replacement therapy or proning. Non-Suvivors have a trend for higher intensity organ support as demonstrated by the higher peak. They also require renal replacement therapy earlier in their stay compared to survivors (Shapiro-Wilk p > 0.05; unpaired t-statistic = 11.899, p < 0.05). The mean peak requirement for renal replacement therapy occured at day –1 for non-survivors and day 8 for survivors.

## Discussion

The survivors, who were younger and had lower severity of disease at presentation (Table 1), had higher PF ratios on admission to ICU. In the majority of survivors, presentation to hospital is later. Non-survivors presented earlier to hospital (p < 0.05), required intensive care therapy earlier in their illness course (p < 0.05), with worse respiratory function but ultimately had similar PF trajectories to survivors once aligned.

**Table 1:** Patient Characteristics demonstrating a lower age and APAPCHE II score in survivors

|  | Non-Survivor | Survivor | P value |
| --- | --- | --- | --- |
|  | (n = 16) | (n = 15) |  |
| **Age** (mean (SD)) | 66.62 (12.45) | 56.07 (6.79) | 0.007 * |
| **Gender** = Male (%) | 14 (87.5) | 13 (86.7) | 1 |
| **APACHE II** (mean (SD)) | 26.19 (4.65) | 20.60 (6.78) | 0.012 * |
| **BMI** (mean (SD)) | 29.22 (11.94) | 27.05 (3.35) | 0.503 |
| **Smoking** (%) |  |  | 0.438 |
| Ex-Smoker | 12 (75.0) | 10 (66.7) |  |
| Never | 3 (18.8) | 5 (33.3) |  |
| Current | 1 (6.2) | 0 (0.0) |  |

Descriptively, in the survivor group, the lymphocyte count recovers near the nadir of the PF ratio and coincides with the peak ferritin count. There doesn’t seem to be a similar trend in ferritin values amongst non-survivors who present with a higher ferritin count and fail to fall.

Organ support and critical care intensity is different in the two groups – survivors rarely required renal replacement therapy prior to the nadir of their PF ratio but non-survivors presented earlier and required greater organ support. The timing of renal replacement therapy is statistically significant; non-survivors required renal support at mean day –8 compared to survivors on mean day +1 (p < 0.05)

ARDSnet defined severe ARDS as a PF ratio < 13.3 kPa (100mmHg) with the PROSEVA trial’s cut-off for proning being a PF ratio < 20 kPa (150mmHg)^9,10^. Taking these cut-offs into consideration, the majority of the survivors presented in this case series do not improve beyond severe ARDS criteria until day 20 – 25 after symptom onset and do not improve beyond PROSEVA’s proning cutoff until much later. In both groups, the amount of organ support required as their disease progresses increases and peaks at the day coinciding with the lowest PF ratio. In survivors this level of support is maintained and stabilises until approximately day 10 post lowest PF ratio. Interestingly, vasopressor usage (as defined as use of any noradrenaline, vasopressin or metaraminol) declines but renal replacement therapy increases post PF ratio trough. Non-survivors require intensive organ support early on in their admission to ICU.

Based on this case series, there is little to differentiate survivors from non-survivors based on respiratory function and lymphocyte count until either recovery or death. Critical care needs, including vasopressors, proning and renal replacement therapy can extend beyond

10 – 15 days after lowest PF ratio in survivors. This is in stark opposition to previous case series who fail to demonstrate trajectories of patients due to non-alignment of disease progression which is the novelty of this case-series.

Limitations of this case series include a small sample size in both survivors and non-survivors with selection bias occurring in the survivor cohort.

## Conclusions

A case series of COVID-19 survivors and non-survivors was presented demonstrating that resolution of severe ARDS/ proning requirements may not occur until beyond day 20 – 30 after symptom onset.

In this series, looking at multiple factors there was little to differentiate survivors from non-survivors upon which to base early prognostication.

Even when faced with multiorgan failure, perseverance until discharge must be advocated as late improvements do occur in survivors.

## Data Availability

No data is shared

